# A foggy minefield: Experiences of regulation among developers of AI and other medical software in the UK, survey and focus group study

**DOI:** 10.1101/2024.08.25.24312551

**Authors:** Henry WW Potts, Paulina Bondaronek, Ana Luísa Neves, Alex Bolotov, Lucie Burgess, Jona Shehu, Gabriella Spinelli, Emanuela Volpi, Austen El-Osta

## Abstract

**Introduction:** Regulation is important for medical software, but advances in software, notably developments in artificial intelligence (AI), are developing quickly. There are concerns that regulatory processes are not keeping up and that there is a need for more pro-innovation approaches.

**Methods:** We conducted a survey (n = 34) and four focus groups to discuss experiences of regulation among UK-based developers.

**Results:** In the survey, 35% agreed/strongly agreed that they were confident in their knowledge of relevant regulation, while 50% agreed/strongly agreed that poor regulation was allowing bad products to come to market. The focus groups identified 10 themes around challenges with current processes: the process of obtaining regulatory approval is uncertain; lack of knowledge about regulatory approval; difficulties in obtaining reliable advice; complexity and slow pace of approvals; difficult to get NHS clinician involvement; process is costly and difficult to fund; implications for competition; international differences; incentives to develop lower classification products; and lack of harmonisation between NHS and MHRA. Respondents’ suggestions for solutions to improve processes fell under four themes: financial and structural support; regulatory collaboration and commissioner involvement; process efficiency and adaptability; and education and guidance.

**Discussion:** Developers are unhappy with the process of regulation for medical software in the UK, finding it confusing and expensive. They feel systems compare poorly to international comparators. Integration between the MHRA system and NHS commissioning is considered poor.

## Introduction

Healthcare is a high reliability industry, with medical errors still a leading cause of mortality (Garrouste-Orgeas et al., 2012). Because of this, there is a high level of regulation: for example, in the training of and professional standards for healthcare practitioners and in the control of pharmaceutical products. Regulation is also needed for medical software, including artificial intelligence (AI). AI is generally regulated as software as a medical device, but introduces its own complexities (Palaniappan et al., 2024). It is critical that medical software and AI products are safe, secure, effective, inclusive and trusted.

While regulation is essential, there are also concerns that an overly complex regulatory landscape might hinder innovation. In the case of digital health, there is much excitement about the possibility of benefit to patients and savings to the healthcare system that could result from adopting digital health solutions. Because digital technologies develop rapidly and more quickly than traditional regulatory systems can keep pace with, there is a call for pro-innovation approaches to regulation (Thierer, 2023; McLean, 2023).

The UK is a global pioneer in digital healthcare and is already progressing evidence-based change in pro-innovation regulatory science (e.g. Medicines & Healthcare Products Regulatory Agency, 2024; Goldacre & Morley, 2022; Department for Science, Innovation & Technology, 2024). Given the strategic importance and rapid technological innovation in this sector, there is an increasing need for a supportive regulatory environment that encourages innovation while ensuring safety, inclusivity, robustness and efficacy. The lack of these conditions hinders the ability of UK businesses to excel, lowers the quality of care that technological progress can deliver, and reduces the global competitiveness of the UK (Spinelli, 2022). The medical regulatory landscape in the UK is complex, particularly given recent developments around the country’s withdrawal from the European Union (Brexit). Medical software in the UK is currently regulated by UK Medical Device Regulations 2002 (which implements EU Directive 93/42/EEC) and is substantially out-of-date compared to the EU, for which new regulations were introduced in 2017 (EU MDR 2017). However, due to the Brexit Windsor Framework, Northern Ireland remains under EU MDR 2017 and thus may also be under the jurisdiction of the recent EU AI Act (European Parliament, 2024). The joint proposal released by the Federal Drug Administration (FDA) in the US, Health Canada, and the MHRA in the UK for healthcare AI regulation and best practices exemplifies the appetite among national regulators for international collaboration (Medicines & Healthcare Products Regulatory Agency, 2021). At time of writing, new regulation in the UK is expected, and the new government elected in July 2024 is expected to make changes in this area.

The regulatory system includes rules, be they laid out in legislation or statutory instruments, but also non-statutory but recommended guidance (e.g. guidelines published by the Medical Devices Co-ordination Group in Europe), best practices (e.g. the NHS Digital Technology Assessment Criteria, DTAC), certifications (e.g. Cyber Essentials), and standards (e.g. ISO 13485). To support businesses with compliance, a variety of services are available, including the provision of knowledge and educational resources, advice, and consultancy. Some of these are provided by the UK government, while others exist as commercial services.

The purpose of regulation is to protect patients and members of the public, and health care systems and civil society more broadly. However, in this sector the users of regulatory systems are those developing medical software and AI, who are often small and medium-sized enterprises (SMEs). In 2020, 85% of the 2,900 businesses in the UK’s digital health sector were small and medium-sized enterprises (SMEs), which accounted for 31% of the sector’s employment and 20% of its turnover (Department of Health and Social Care, Office for Life Sciences & Department for Business, Energy & Industrial Strategy, 2021; Department of Health and Social Care, Office for Life Sciences & Department for Business, Energy & Industrial Strategy, 2024). As with any service, it is important to listen to users’ experiences to deliver user-centred services. In this study, we investigated the perceptions and experiences of developers of medical software, including AI, of regulation.

## Methods

We conducted a survey and four focus groups to discuss experiences of regulation among UK-based developers. The survey was developed based on expert knowledge in the team and our prior work with innovators in the field. The survey and focus groups were centred on three key themes: access to regulatory knowledge and expertise, innovation barriers in the health tech market, and possible solutions to enhance the health tech ecosystem. The ethics committee of the University of Westminster Research gave ethical approval for this work (#ETH2324-1792).

Both the survey and the focus group invitations were addressed to developers of medical software, including AI. The focus groups were also open to those working in regulation. We allowed respondents to self-identify.

The survey used a mix of multiple choice and open-ended questions. It was hosted on the JISC platform. The survey and focus groups were promoted through social media (LinkedIn, X and BlueSky, with most engagement taking place on LinkedIn) and the personal networks of the research team. We asked people to forward the survey and focus group invitations to any relevant individuals or forums, i.e. snowball sampling. Data collection took place May-July 2024.

We held four focus groups. There were two in person events in London in June 2024: the first involved one focus group, while the second had two focus groups in parallel. An additional focus group was held online, over MS Teams, in July 2024. The in-person groups (4-5 participants each) were roughly 90 minutes long, with a break in the middle. The online group (4 participants) was shorter, around 45 minutes in total, with no break. We did not collect demographic information on focus group participants. Some focus group participants had previously filled in the survey. Focus groups were preceded by a presentation on the regulatory landscape. Two broad questions were asked: (1) what are the challenges to the regulatory system, and (2) what would be some solutions to those challenges?

The focus groups were not recorded, but we used 1-3 note-takers per focus group to capture key insights from the discussions. Note-takers were mainly silent, but would also ask questions for clarification. Members of the research team also participated in discussions on some occasions. We conducted a thematic analysis (Fugard & Potts, 2019) of the focus groups notes: HWWP did the initial analysis, which was then separately reviewed by PB and ANL.

## Results

### Survey

We received a total of 34 responses to the survey, though it was not possible to calculate a response rate as we do not know how many people saw the survey on social media or via other channels.

The majority of our respondents (62%) were in micro-businesses (under 10 employees) and were making medical software with AI. 41% were in market. Most were selling directly to the NHS or other healthcare providers (85%), with half selling to other businesses. Most were operating in England (85%), with over a quarter operating in Wales (29%), the European Union (26%), the US (29%) and the rest of the world (i.e., not the UK, EU or US; 32%). Professional backgrounds were varied (modal answers were product design/development and project management, 41% each).

Only a minority were confident in their knowledge of relevant regulation (35% agree or strongly agree). Half felt poor regulation was allowing bad products to come to market (50%).

In terms of specific challenges faced, the main ones were with NHS commissioning (62%) and selecting and collecting evidence that the regulator accepts as sufficient and relevant (53%). Over a third also mentioned: carrying out evaluation studies (47%), research ethics approval (41%), data access and sharing (41%), and data ownership (35%).

Half knew where to find expertise and resources to help (50%), with the main source of help being from professional consultancy (62%), while over a third (44%) also said other companies who have previously experienced the regulatory process, government agencies (41%) or their own in-house expertise (41%). Affordability was an issue, with over half disagreeing with the proposition that getting help on regulatory matters was affordable (59%).

About two thirds also disagreed that they were confident about their knowledge of upcoming changes in the regulatory process (65%).

More agreed than disagreed that the current processes require an appropriate amount of time and resources (39% versus 32% respectively), but the group was fairly evenly split.

About two thirds (65%) disagreed that the regulatory approval process was clear about the evidence required and the methods to gather them. Half disagreed with the assertion that finding a clinical partner was easy (50%) and nearly two thirds (62%) found doing so expensive.

The relationship between the current regulatory approval and NHS commissioning was seen as poor. Over half characterised the two as uncoupled and making the processes more complex (59%) and as inconsistent with each other (53%).

### Focus groups Challenges

We identified 10 themes in terms of challenges faced by innovators: see Table 3.

**Table 1:**
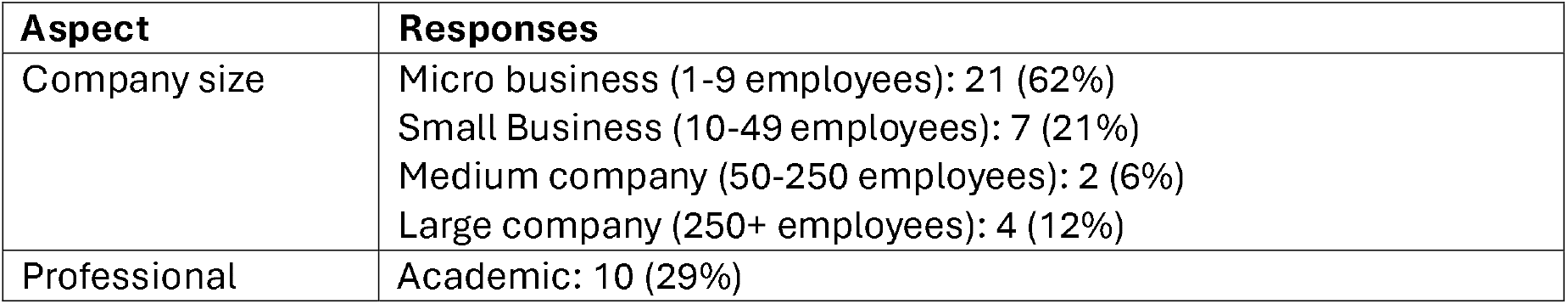
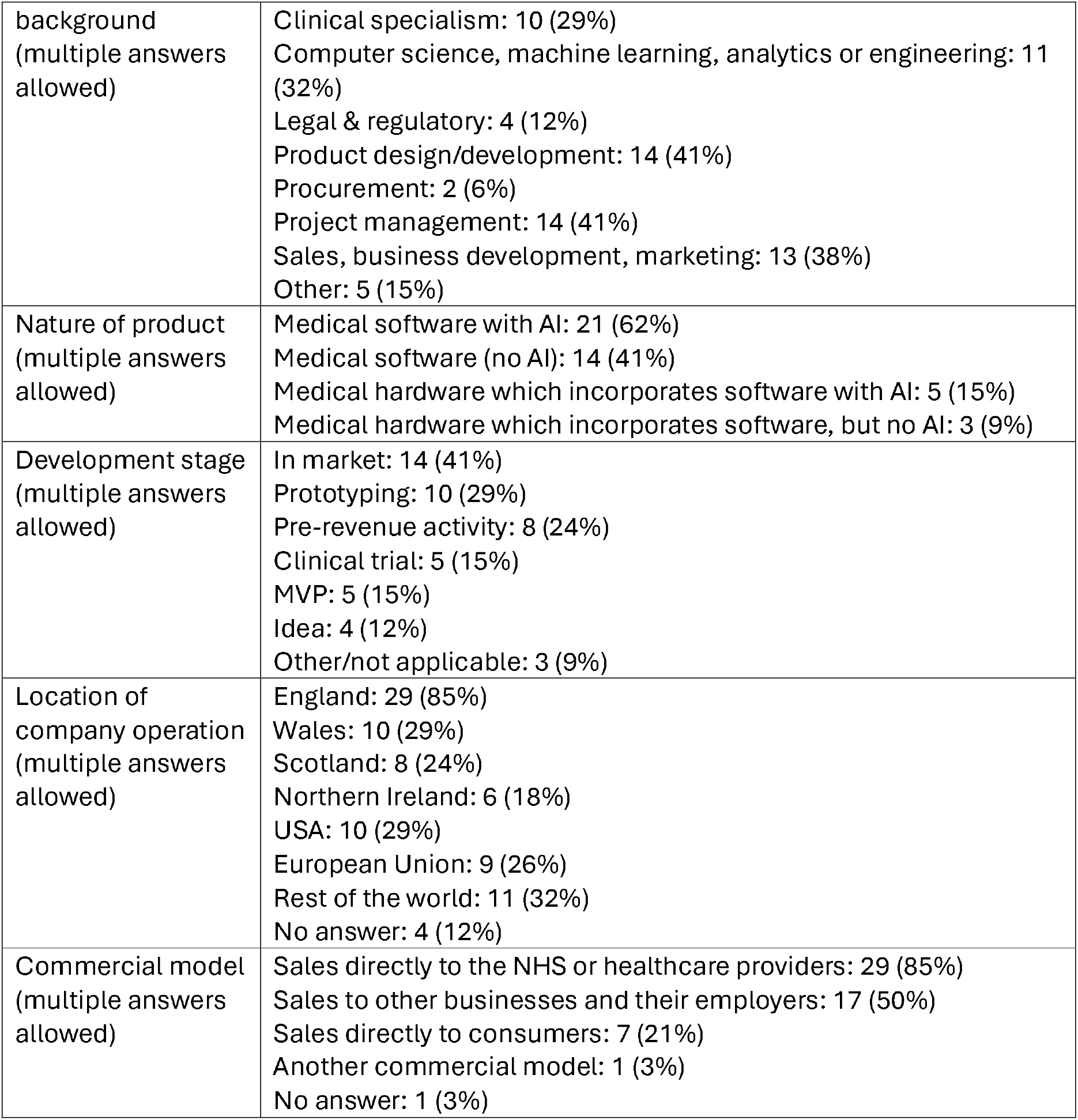
Description of respondents.

**Table 2:**
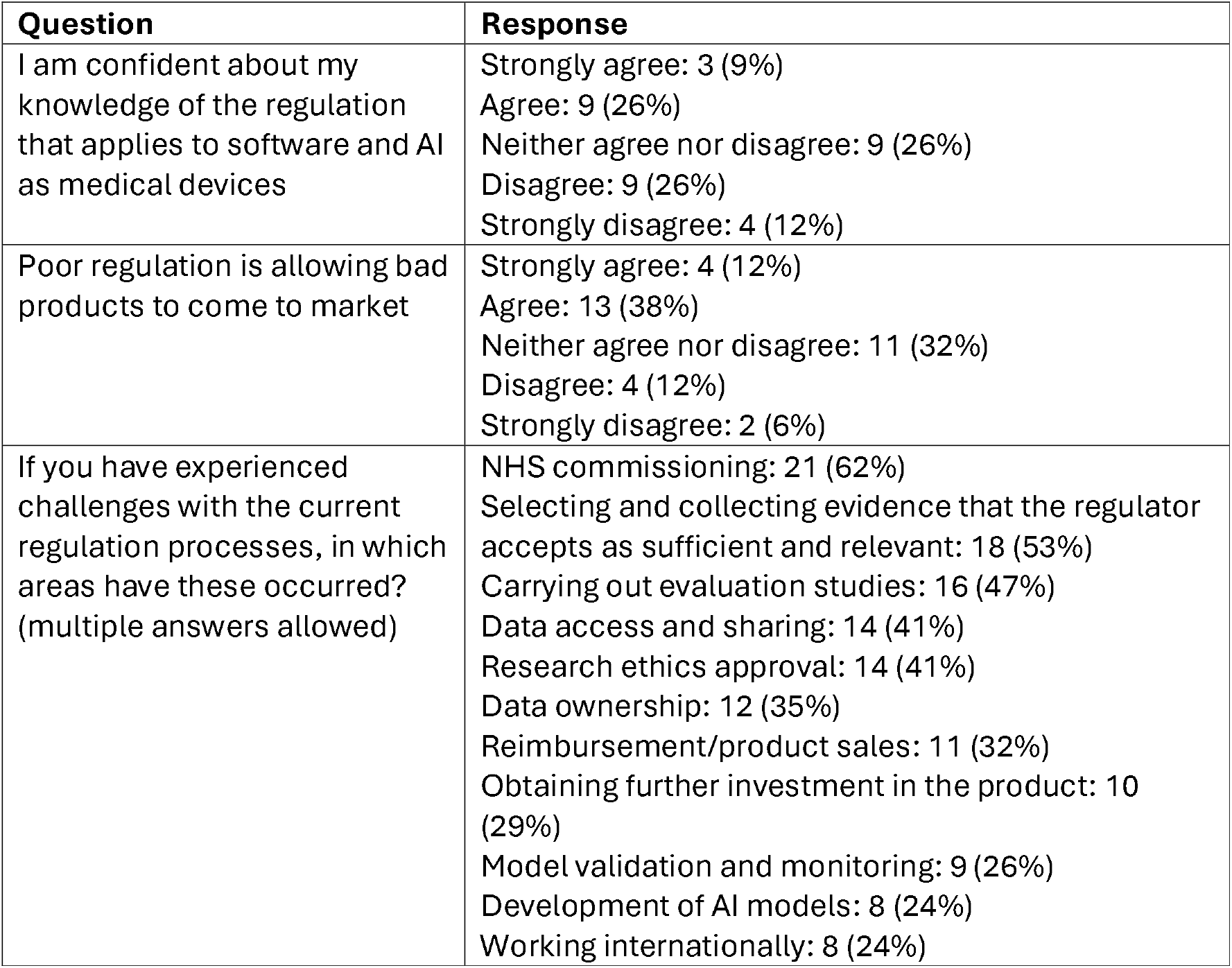
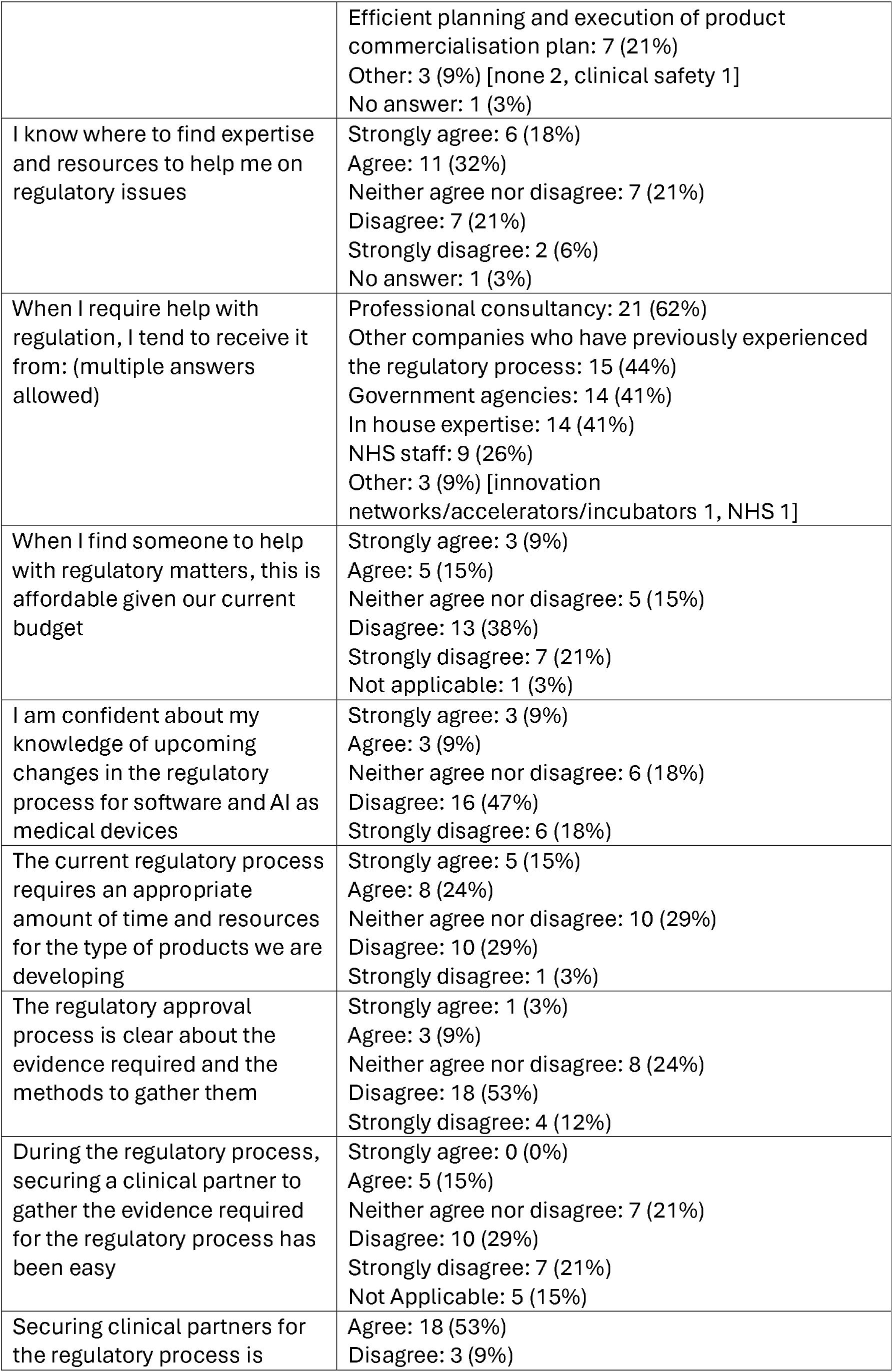
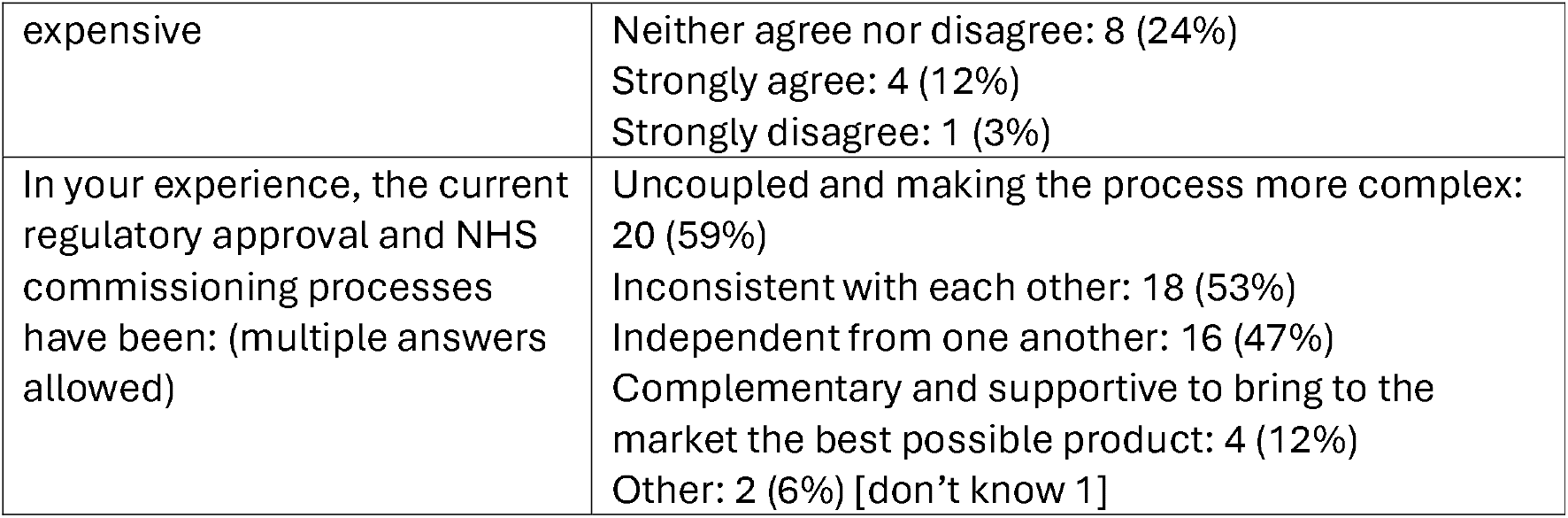
Experiences of regulation.

**Table 3:**
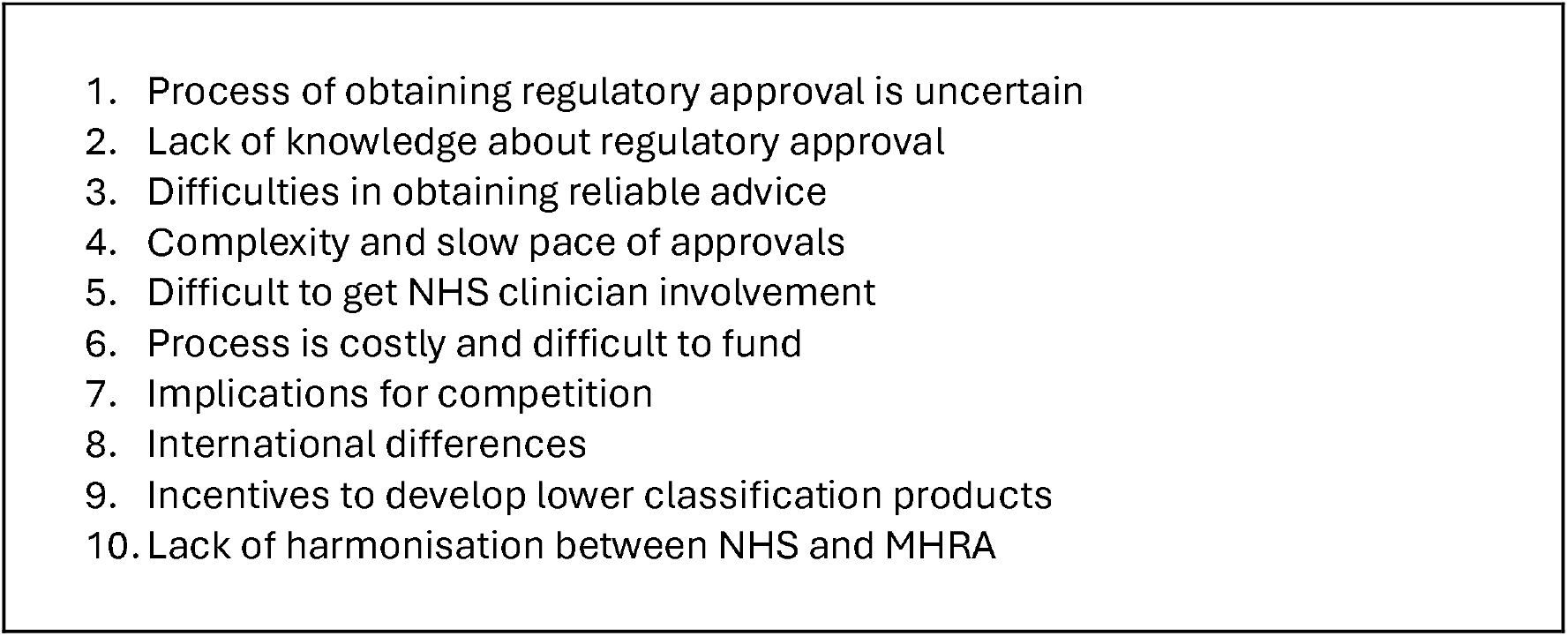
Challenges identified by innovators with regulation.

#### Process of obtaining regulatory approval is uncertain

Participants described the process of obtaining regulatory approval as being uncertain. One participant said, “It feels like walking through a foggy, unmarked minefield”. What MHRA classification (if any) will apply to one’s product was unclear, and while decision trees exist to answer this question, the experience of trying to follow one was likened to entering a “maze” or going down a “rabbit hole”. Participants felt there was a lack of guidance available on the process. There was frustration that the MHRA refused to provide clear determinations, unlike the FDA and regulators in Europe. Areas that were mentioned as being particularly complex/borderline were mental health apps, screening devices, and risk calculators.

The process being unclear means, in turn, that costs and timelines are unclear, making planning difficult. Likewise, this makes the return on investment of activities unquantifiable. There were further specific areas of uncertainty expressed, including concern about the ambiguity of ethical processes in AI model development and a perceived lack of quality standards for training data and algorithmic architecture.

##### Lack of knowledge about regulatory approval

Participants described a lack of knowledge about the regulatory system among those applying for approval. This included companies not realising what the costs of obtaining regulatory approval will be, and also that investors (existing or potential) do not realise the costs or understand the scope and importance of regulation.

##### Difficulties in obtaining reliable advice

There was uncertainty around who can provide advice on the regulation journey. Precious time is spent waiting for answers. Advice obtained, including from paid consultants, was perceived to often not be reliable, being unclear or contradictory. There was criticism of the role of the MHRA, which was described as being insufficiently proactive in engaging with potential applicants. Answers obtained from the MHRA were felt to be poor and they were criticised for not giving definitive answers (with the US FDA considered to do better), although participants also acknowledged that innovators’ ideas can evolve rapidly, which makes it difficult for a regulator to provide determinative answers.

##### Complexity and slow pace of approvals

The process of obtaining evidence for regulatory approval was described as being complex and slow, with access to data a particular challenge. Participants reported faster innovation timelines outside the UK.

##### Difficult to get NHS clinician involvement

Getting NHS clinician involvement was described as being difficult. There was an awareness of why this should be the case and of how busy clinicians are.

##### Process is costly and difficult to fund

Because of the aforementioned factors and because of the nature of clinical research, the whole process of obtaining regulatory approval, including in terms of clinical studies for evidence generation, is costly. Funding is needed from somewhere. While it is possible to apply for some public funding for research, this process is slow in terms of waiting for appropriate calls, and waits for a decision.

##### Implications for competition

These aforementioned costs have important implications for competition. Costs are the same irrespective of company size, which is a particular challenge for the many SMEs in digital health, making it harder for them to function and compete, while larger companies can more easily absorb them. Larger companies were perceived as having in-house teams with regulatory skills and less of a need to use consultancies.

##### International differences

The FDA in the US was seen as offering a better experience, but it was recognised that the FDA has substantially greater funding. The US was seen as an important market, with more investment perceived to be available for digital health start-ups.

International differences in regulation were unwelcome, with companies having to go through separate processes in different markets. Given the costs of obtaining regulatory approval, there was an incentive to pick a bigger market, like the US, and just to obtain approvals there. More harmonisation was desired. It was felt to be bad for the UK economy if we do not help companies with the regulatory challenges they face in other markets because otherwise UK companies will struggle to export.

##### Incentives to develop lower classification products

The current system was seen to incentivise game-playing. There was pressure to aim for a lower classification and to develop products that did as much as possible without tipping into a higher classification. There was a disincentive to develop new features for a product that would lead to a higher classification.

##### Lack of harmonisation between NHS and MHRA

Participants talked about a lack of harmonisation between the MHRA’s and the NHS’s decisions. Achieving regulatory approval from the MHRA and the process of being commissioned by a part of the NHS are disconnected processes. Participants wanted a more integrated experience. Likewise, it was felt that NICE and MHRA were not coordinated.

### Solutions

We identified key themes in terms of possible solutions: (1) Financial and structural support, (2) Regulatory collaboration and commissioner involvement, (3) Process efficiency and adaptability, and (4) Education and guidance.

#### Financial and structural support

Participants wanted more financial support for innovators – be that subsidising the journey through the regulatory process, more public money for evaluation studies, or the use of public money rather than venture capital to fund innovation – and more support generally. It was felt that digital health products are a public good, so there should be more publicly funded support. It was suggested that large companies should be taxed more to pay for more support for innovators in SMEs.

The idea of more support services was welcomed, which participants felt could be paid for by the government or by innovators. That service, participants said, should have formal connections to the MHRA and/or NHS and/or NICE. This was because of a strong desire for advice that could be considered authoritative, because there is a need for certainty and accuracy in the advice.

Innovators want to talk to a human. They talked of a concierge or navigator service, or of having an assigned case worker at the regulator. However, other tools would also be useful. For example, there were suggestions to develop a decision tool (one participant made a comparison to the NHS Health Research Authority research ethics tool; NHS Health Research Authority, n.d.), a flow chart or road map, application templates, or a large language model-based chatbot, although the question of whether the latter would be reliable enough was raised.

Desired advice to be provided included an estimate of costs as this is an important item in the budget to present to investors and venture capital. There was also the suggestion for an observatory tracking new developments in AI.

#### Regulatory collaboration and commissioner involvement

Participants wanted the government and regulators to work more collaboratively with innovators. One participant put it that, “Regulators need to come on a journey with us.” There was a suggestion for a hybrid public/private body sitting between the NHS and innovators, or for the NHS and SME to share the risks of developing innovative products.

Generally, there was a desire for the regulators to be more active and provide more input. This came with the recognition that the MHRA should be better funded.

There was discussion around the relationship between regulation and commissioning. Participants saw a value in there being greater consistency in NHS practice and better alignment between NHS trusts/integrated care systems at one level and NHS England at the top. Several participants suggested the system for developing innovation in digital health should be more led by needs. For example, it was proposed that commissioners should identify areas that need solutions and then commission solutions, with that commissioning process then including help with regulatory processes.

Various comments suggested the MHRA should look to the FDA as a model. For example, participants suggested that the FDA gives clearer feedback on classifications and will guide innovators to appropriate programmes. Better alignment with other nations, particularly US and EU, was desired, be that simply the UK copying US or EU regulations, or creating a standardised process to go from being validated in the UK to being validated in the US.

#### Process efficiency and adaptability

Participants wanted processes to be simpler, clearer, faster and cheaper. That was tempered with a recognition that innovators’ products can change, which makes it difficult to have a simple regulation journey, and it was acknowledged that some companies try to cheat the system. Participants wanted a better system to handle when new features are added to an existing product, and specified that over-demanding regulatory processes required for product updates stifle innovation.

Several participants suggested some sort of system that has different evidence requirements at different stages of development, such that there is a lighter touch while a product is being developed and initial evidence is being gathered. Similarly, it was suggested that the NHS should support start-ups at the lowest possible classification and allow them to collect data with that version of a product, and then they can move up classification later if needed, as opposed to the current system that was seen as being risk averse and wanting higher classifications from the start. One participant mooted the idea of recognising a “minimum viable medical product” and the need to support companies to generate revenue while they were still collecting evidence.

Various initiatives to speed up the regulatory process were mentioned as being welcome: the MHRA AI Airlock, the Information Commissioner’s Office (ICO) sandbox, and NICE Early Value Assessments.

#### Education and guidance

The idea of having educational offerings on regulation was discussed, with various specific ideas suggested. Face-to-face education was preferred, but the use of MOOCs (massive open online courses) was also proposed. Teaching that was practical, contextualised, and as brief as possible was wanted. There was a particular desire for investors/venture capital to be better educated around regulation.

## Discussion

Responders lacked confidence in their knowledge of regulation and future regulatory changes. They found the system confusing and mysterious. They could struggle to find affordable help, with consultancy being the main source. They did not trust the advice they received.

They were critical of processes. Slightly more survey respondents thought processes required an appropriate amount of time and resource as not, but respondents felt the current system allowed bad products to come to market and that they did not link well with NHS commissioning. They felt processes were unclear about the evidence required. Getting a clinical partner was reported to be difficult. Other specific challenges included carrying out evaluation studies, obtaining ethics approval for studies (compare Yang & Potts, 2024) and issues over data (access, governance, ownership).

While academic literature focuses on barriers to innovation from an economic perspective and the importance of regulations from a patient safety perspective, the views of innovators is often neglected or considered anecdotal. This paper articulates consistent threads that have emerged through bottom-up engagement to voice concerns, that, to date, is spoken of but often ignored in academic publications, despite the considerable economic power and innovation potential that SMEs deliver in the UK healthcare technology market.

We should not presume that everything innovators want is practical. There are other stakeholders in the process, whose needs should be balanced (Zhou & Gattinger, 2024). It is no surprise that innovators want costs burdened by someone else, and of course innovation activities are already subsidised by the state (Mazzucato, 2013). However, it does appear reasonable to conclude that there are areas for improvement. There are questions for government around how much they wish to support innovation in this area. Participants saw the value of alignment with other countries, but also there was a sense of poor alignment within the UK between MHRA and NHS, particularly NHS commissioning. Better coordination might be possible at minimal cost.

Innovators struggled with the lack of certainty around regulatory decisions. Developers bemoaned the difficulty of getting definitive answers on what regulation they will need, but the difficulty of that task should not be underestimated. There was recognition that companies’ ideas frequently evolve given the nature of the technology and the incremental understanding of the clinical challenge/context. Participants’ desire for an authoritative answer early in a development cycle may not be entirely achievable. However, more emphasis on defining products/systems at an early stage, e.g. through requirements engineering (Spinelli, 2022) or the use of logic models (Potts et al., 2021), may help.

## Conclusion

Innovators are unhappy with the process of regulation for medical software (including AI) in the UK, finding it confusing and expensive. They feel systems compare poorly to international comparators. Integration between the MHRA system and NHS commissioning is also considered poor.

## Data Availability

All quantitative data produced in the present study are available upon reasonable request to the authors.

## Declarations

## Acknowledgements

Thanks to the participants for their time. Thanks to the other members of the RADIANT team, including Aaron Kandola, Allan Tucker, Amedeo Chiribiri, Claire McCallum, and Justine Kenyon. Prof Paul Wallace was also part of the team before his death in February 2024. We dedicate our work to his memory.

## Contributions

Everyone designed the study. EV led on obtaining ethics approval. AB led on survey implementation. HWWP/GS/CM/LB/AB/ALN/AEO designed the survey and workshop activities. Everyone helped disseminate the survey and call for focus group participants. AB/EV/JS organised the focus groups. LB/AB/HWWP/GS/CM chaired the focus group workshops. HWWP/JS/PB took notes. HP did the quantitative analysis. HWWP/ALN/PB did the qualitative analysis. HWWP wrote the first draft. All authors reviewed and inputted into the manuscript.

